# Altered interoception in schizophrenia and its role in subjective emotional experience

**DOI:** 10.64898/2026.09.23.26363817

**Authors:** Beier Yao, Kathryn Eve Lewandowski, Mei-Hua Hall

**Affiliations:** Schizophrenia and Bipolar Disorder Program, McLean Hospital, Belmont, MA; Department of Psychiatry, Harvard Medical School, Boston, MA; Department of Psychiatry, Mass General Brigham, Boston, MA

**Keywords:** heartbeat-evoked potential, EEG, ECG, heartrate, physiology, arousal

## Abstract

**Background:** Interoception refers to the processing, integration, interpretation, and regulation of bodily signals by the brain. It is crucial to motivational and affective functioning such as subjective experience of arousal (i.e., intensity of emotion). Previous studies found altered self-report of arousal in people with schizophrenia, but the underlying biological mechanism is unclear. This study aimed to investigate interoceptive processing and emotional experience in schizophrenia by triangulating self-report, cardiac dynamics, and heartbeat-evoked potential (HEP; an event-related potential that reflects cortical processing of heartbeats).

**Methods:** A total of 39 participants with schizophrenia spectrum disorders (SZ; 36% female) and 31 healthy controls (HC; 35% female) reported subjective arousal level when viewing images of varying emotional intensity while electroencephalogram and electrocardiogram were being recorded.

**Results:** We found that intense images led to larger heartrate decelerations and higher subjective arousal ratings in HC, but did not modulate heartrate changes in SZ and were less predictive of their subjective ratings. Subjective ratings were predictive of cardiac dynamics in HC, but not in SZ. SZ showed a less negative HEP amplitude than HC when viewing less intense images. Lastly, SZ with less severe positive and negative symptoms exhibited cardiac dynamics more similar to those of HC.

**Conclusions:** These findings suggest both reduced cardiac reactivity to emotional stimuli and a disconnect between interoceptive afferent signals and cortical processing, leading to different subjective emotional experience in people with schizophrenia. Studying interoception using a multimodal approach may reveal important insights into the underlying mechanisms of altered emotional experience in schizophrenia.

## Introduction

Interoception refers to the processing, integration, interpretation, and regulation of bodily signals by the central nervous system. There is growing evidence that internal signals from the body can influence a wide range of brain functions, including basic perception, processing speed, reasoning, emotion, motivation, and our sense of self (1–4). Therefore, interoception could be a common mechanism affecting many different symptoms and impairments in schizophrenia. Though experimental evidence is limited, there is nevertheless a large body of peripheral evidence suggesting altered interoception in schizophrenia (5).

One important function that interoception serves is to enable the subjective experience of arousal (i.e., intensity of emotion) (6,7). One commonly used physiological index of arousal is momentary heartrate deceleration (8). It has been found that individuals with better interoceptive ability (as measured by their ability to count their own heart beats) demonstrate a higher level of correspondence between self-report arousal and heartrate deceleration than those with poor interoceptive ability do (8). Moreover, those showing a stronger correspondence between subjective and physiological arousal also exhibit a stronger intrinsic connectivity within the brain network posited to support interoception (9), demonstrating a direct brain-body connection. Therefore, the correspondence between subjective and physiological arousal could serve as a useful proxy of interoception, especially when examining its impact on emotional experience.

Most existing studies have only examined physiological and subjective arousal separately in persons with schizophrenia. A meta-analysis revealed that persons with schizophrenia and healthy participants reported similar levels of subjective arousal in response to evocative stimuli (both pleasant and unpleasant), but persons with schizophrenia reported higher arousal than healthy controls in response to neutral stimuli (10). Very few studies have examined physiological measures of arousal in persons with schizophrenia, and found a mix of similar and different patterns of heartrate changes compared with healthy participants (11–13). Without examining subjective and physiological measures simultaneously, it is unclear whether self-reports of increased arousal reflect a response bias (and thus a disconnect with bodily signals) in persons with schizophrenia, or an abnormally high bodily response to neutral stimuli (and thus an accurate report of an elevated arousal state at baseline).

A more recent measure of interoceptive processing is the Heartbeat Evoked Potential (HEP) - an event-related potential in the brain that is time-locked to heartbeats, which purportedly reflects cortical processing of cardiac signals regardless of whether heartbeats were consciously perceived (14). One advantage of this index is that the amplitude of HEP can be modulated by attention, interoceptive ability, psychiatric conditions, and especially arousal state (14). There is preliminary evidence of smaller HEP amplitude at rest (15,16) and altered attention modulation of HEP (17) in schizophrenia. To our knowledge, no prior study has examined arousal modulation of HEP in schizophrenia, and very few studies have examined HEP during emotional image viewing in any population. This makes the present paradigm a novel test of whether schizophrenia is associated with altered cortical processing of cardiac afferent signals during emotional arousal.

The current study aimed to investigate interoception in schizophrenia by examining the correspondence between physiological and subjective arousal states, as well as the interactions between cardiac signals and central processing. We hypothesized that relative to controls, participants with schizophrenia would have less correspondence (i.e., a greater disconnect) between momentary heartrate deceleration and self-reported arousal level. We also hypothesized that participants with schizophrenia would show altered HEP amplitude and altered arousal modulation of HEP. Due to the lack of prior studies, we did not have specific hypothesis on symptom correlations and conducted exploratory analyses on the relationships between interoception and symptom severity. Findings from this study will add to our knowledge of interoception in schizophrenia and the biological bases of subjective emotional experience.

## Methods and Materials

### Participants

Participants with schizophrenia-spectrum disorders (SZ) were recruited through psychiatric services at McLean Hospital, existing research registry and subject pools, and community and online advertisements. Healthy control participants (HC) were recruited through online advertisements and existing research registry. All participants were between the ages of 18-60 and fluent in English. Diagnoses were based on the Structured Clinical Interview for DSM-5 (SCID-5)(18) conducted by trained personnel, incorporating information from medical records when necessary. See supplemental methods for exclusion criteria. The final sample included 39 SZ (19 with schizophrenia disorder, 19 with schizoaffective disorder, 1 with delusional disorder), and 31 HC (1 with past alcohol use disorder and 1 with past generalized anxiety disorder). For EEG analysis, 4 additional SZ participants were excluded due to poor data quality overall (2 due to hairstyle-related artifacts, 2 due to frequent drifting in waveforms that cannot be corrected). All participants provided written informed consent and were compensated for their participation. The Mass General Brigham Institutional Review Board approved the study.

### Assessments

For SZ, we assessed psychotic and mood symptoms within one month of study visit using the Scale for the Assessment of Positive Symptoms (SAPS)(19), Brief Negative Symptom Scale (BNSS)(20), Young Mania Rating Scale (YMRS)(21), and Montgomery-Asberg Depression Rating Scale (MADRS)(22). Duration of illness was estimated from participants’ onset of first psychotic symptom. Community functioning was assessed using a version of Multnomah Community Ability Scale (MCAS)(23) excluding items on symptom severity(24). For participants taking antipsychotic medication, we calculated chlorpromazine (CPZ) equivalents(25–27). All native speakers of English completed the North American Adult Reading Test (NAART)(28) to estimate premorbid IQ. See supplemental methods for self-report scales.

### Task and Procedure

Participants viewed and rated images while their electrocardiogram (ECG) and electroencephalogram (EEG) were recorded. Ninety-six photos were selected from the International Affective Picture System (IAPS)(29) and used to induce heartrate changes and corresponding subjective feelings of arousal. The photos were selected based on normative ratings of valence and arousal, and contained 32 pleasant, 32 neutral, and 32 unpleasant images(30). Each trial started with a small white fixation dot (diameter: 10 pixel) in the center of a grey screen (with the same brightness as the photos) for a jittered interval of 7-9 seconds (Figure 1). Then participants viewed a photo for 6 seconds. Following the disappearance of the photo, participants were asked to rate their experience while viewing the image on valence and arousal using the self-assessment manikin (SAM)(31), a nonverbal rating scale that uses figure drawings to represent the valence and arousal dimensions of affective experience. We used the SAM because it was the scale used to develop normative ratings for all IAPS photos(29). Ratings along the SAM corresponds to 1-9 numerically (valence: 1 = most unpleasant, 9 = most pleasant; arousal: 1 = lowest arousal, 9 = highest arousal). See supplemental methods for more details on the task and experimental apparatus and setup.

**Figure 1.**
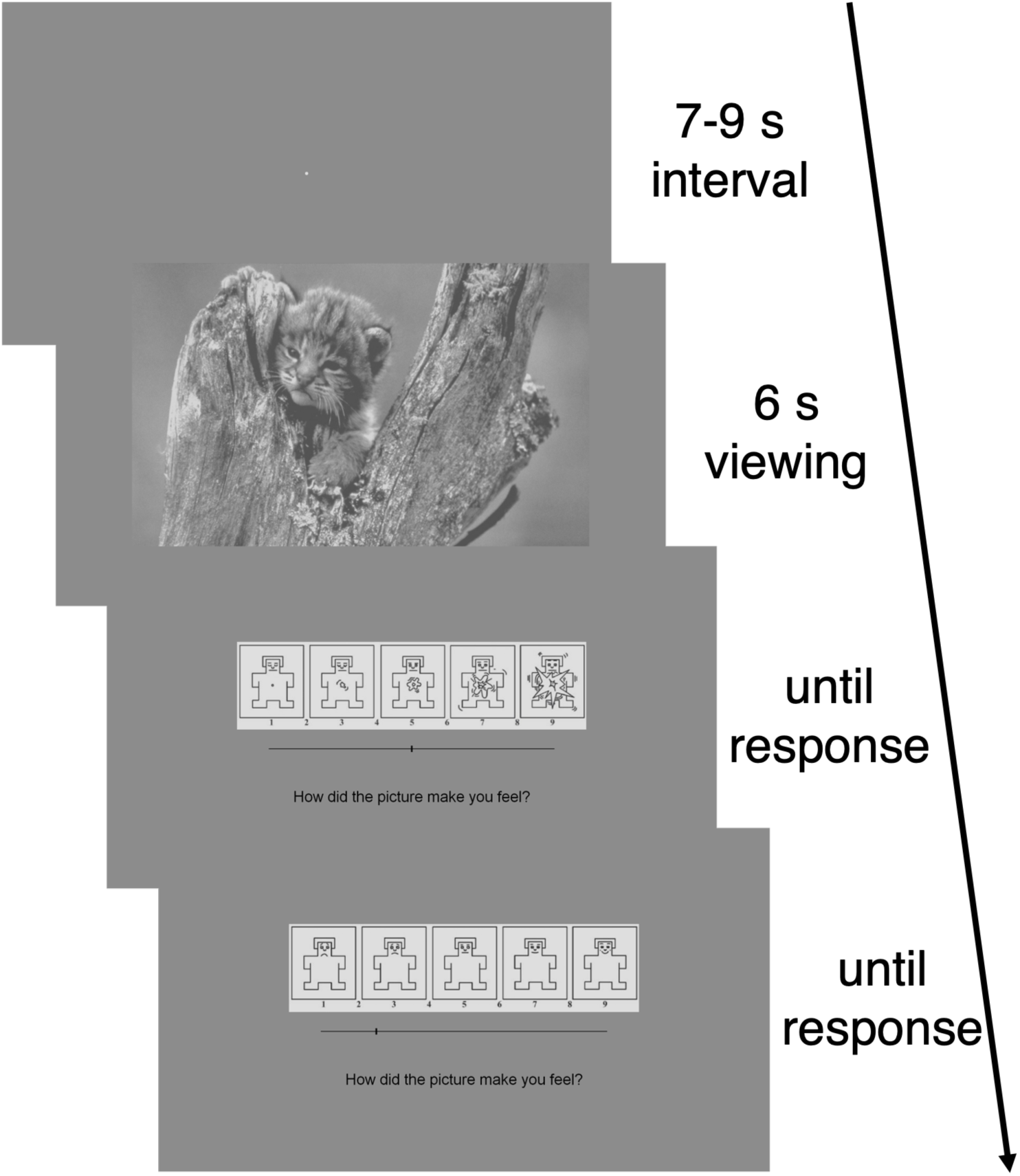
Task procedure. Each trial started with a fixation period for a jittered interval of 7-9 seconds. Then participants viewed a photo for 6 seconds and rated their feelings on arousal and valence dimensions.

### Data Analysis

See supplemental methods for detailed preprocessing steps of ECG and EEG data.

#### IBI analysis

We calculated inter-beat-intervals (IBI) as the time difference between two consecutive R-peaks. For event-related heartrate analysis, we followed established procedures of assessing phasic heartrate changes accompanying stimulus processing(32). For each trial, six IBIs around stimulus onset were extracted for analysis: IBI 0 was the interval concurrent with stimulus onset, preceded by IBI – 1 and followed by IBIs 1, 2, 3, and 4. The second IBI before stimulus onset (IBI - 2) was used as the baseline IBI and deducted from all following IBIs to derive relative changes in IBI after image onset.

#### HEP analysis

We grouped trials showing positive and negative images as the high arousal (HA) condition (mean normative arousal rating = 5.7), and trials showing neutral images as the low arousal (LA) condition (mean normative arousal rating = 3.6). According to a meta-analysis, HEP is typically observed in fronto-central electrodes(14). Therefore, we focused our analysis on Fz, FCz, and Cz channels. To examine potential lateralization effects, we also calculated means of channels within sections(33): F-L = mean of F1, F3, F5; F-R = mean of F2, F4, F6; FC-L = mean of FC1, FC3, FC5; FC-R = mean of FC2, FC4, FC6; C-L = mean of C1, C3, C5; C-R = mean of C2, C4, C6. Based on the meta-analysis evidence that arousal effects on HEP are the strongest approximately 200-300 ms after R-peak(14) and a visual inspection of the grand average waveforms from the HC group, we used 200-330 ms window after R-peak to identify HEP peak for all participants. Because HC provide the reference pattern for typical cortical processing of cardiac signals, we defined the scoring window and polarity from the HC normative response and then applied the same scoring rule to both groups. The HC waveform in our sample showed a clear condition-sensitive response in the 200–330 ms window, with a negative peak in the LA condition and a positive peak in the HA condition (see Figure S1), consistent with prior IAPS-based HEP work (34). Therefore, we selected the highest positive peak in the 200-330 ms window after the R-peak to calculate HEP peak amplitude for the HA condition, and the lowest negative peak for the LA condition. The significant LA-versus-HA condition effects observed across groups subsequently confirmed that this polarity-based scoring captured the expected arousal modulation in each group (see supplemental results). Peak amplitude from both conditions were extracted for statistical analysis.

### Statistical Analysis

We used independent *t* tests and *χ^2^* tests to compare groups on demographic and psychosocial characteristics. For these analyses, we used SPSS Statistics version 28.0 (IBM, Armonk, NY). We then constructed a series of multilevel linear regression models with restricted maximum likelihood estimation for analyses on group differences in subjective rating, heartrate changes, relationships between heartrate changes and HEP amplitude, and symptom moderation effects on heartrate changes. See supplemental methods for more details on these regression models. For group differences in HEP amplitude, we conducted mixed Analysis of Variance (ANOVA) tests. For these analyses, we used R Statistical Software version 4.5.0 (R Core Team, Vienna, Austria). Because we found no effects of age, sex, body mass index (BMI), and CPZ equivalent on baseline heartrate or HEP amplitude (see supplemental results), we did not include these as covariates in analyses to preserve statistical power.

First, we examined the correspondence between IAPS normative arousal ratings and participants’ subjective arousal ratings, and found that the subjective rating was more closely aligned with the normative rating in HC than in SZ (see supplemental methods, results, & Figure S2 for details).

#### Heartrate changes

To assess the effect of IAPS images on heartrate changes, we constructed a multilevel model predicting relative changes in IBI using a combination of time, group, and IAPS normative ratings. Because previous studies found that heartrate changes while viewing IAPS images were non-linear(30,35), we included the quadratic term for time: time^2^. Because normative valence rating may modulate heartrate changes too(30,35), we included both IAPS normative arousal and valence rating as predictors. To account for general inter-individual and inter-trial differences in heartrate changes, we included a random intercept for subject and a random slope of time for trial within subject (Model 1).

To assess the correspondence between subjective arousal rating and transient heartrate changes, we constructed a second model (Model 2) where IAPS normative ratings were substituted with participants’ subjective ratings. The random effects were identical to Model 1.

#### HEP amplitude

To assess the effect of IAPS images on HEP amplitude, we conducted mixed ANOVA tests on peak amplitude from Fz, FCz, Cz, F-L, F-R, FC-L, FC-R, C-L, and C-R, with condition (LA vs. HA) as the within-subject variable and group (SZ vs. HC) as the between-subjects variable. Our main effect of interest was the group × condition interaction, given that HEP amplitudes were mostly positive in HA and mostly negative in LA condition. For significant group × condition interactions, we followed up with one-way ANOVA tests to assess group effects within condition and condition effects within group.

#### Relationship between heartrate changes and HEP amplitude

To assess whether HEP amplitude differences are due to cardiac dynamic differences, we first calculated the mean IBI change series across trials within HA and LA conditions for each participant. For channels with a significant group × condition effect, we conducted a multilevel regression model predicting relative changes in IBI using time, group, and HEP amplitude.

#### Symptom associations

We explored whether the pattern of heartrate changes varied as a function of symptom severity in SZ. We examined the effects of positive and negative symptoms by including sum of SAPS global ratings and BNSS total scores in two separate models. In each model, we included the corresponding clinical measure as moderators of main and interaction effects. We also explored associations between HEP amplitude and clinical symptom severity (see supplemental methods & results).

## Results

### Participant Characteristics

The two groups did not differ on age, sex, and parental education (Table 1). Participants self-identified as Asian or Indian (18.6%), Black or African American (8.6%), Hispanic (10.0%), Multiracial (8.6%), and white (61.4%). The two groups did not differ on the proportion of white (*χ^2^* = 2.66, *p* = .63) or Hispanic (*χ^2^* = 0.006, *p* = 1.0) participants. SZ has fewer years of education, lower premorbid IQ, higher BMI, and faster baseline heartrate than HC. In the SZ group, 84.6% of participants were prescribed antipsychotic medication.

**Table 1.** Demographic and clinical information.

|  | SZ (*n* = 39) | HC (*n* = 31) |  |  |
| --- | --- | --- | --- | --- |
|  | Mean (SD) | Mean (SD) | Statistics | *p* |
| Age (Years) | 33.99 (9.69) | 33.09 (10.44) | *t* = 0.37 | .71 |
| Sex (Female/Male) | 14 / 25 | 11 / 20 | $\chi^2$ = .001 | .97 |
| Education (Years) | 14.82 (1.96) | 16.34 (1.66) | *t* = -3.51 | < .001 |
| Parental education<sup>a</sup> | 5.92 (1.20) | 5.61 (1.61) | *t* = 0.93 | .36 |
| NAART FSIQ | 107.39 (8.52) | 111.71 (7.33) | *t* = -2.18 | .033 |
| BMI | 29.12 (7.16) | 24.69 (3.69) | *t* = 3.34 | .001 |
| Baseline heartrate | 83.22 (13.02) | 69.56 (9.38) | *t* = 5.10 | < .001 |
| IAS | 86.44 (14.19) | 89.81 (10.28) | *t* = -1.11 | .27 |
| IATS | 43.72 (15.48) | 40.42 (19.88) | *t* = 0.78 | .44 |
| ASI3 | 24.72 (16.80) | 7.42 (7.22) | *t* = 5.79 | < .001 |
| TAS-20 | 53.52 (13.03) | 38.39 (8.88) | *t* = 5.76 | < .001 |
| IPASE | 124.04 (48.99) | 69.35 (19.74) | *t* = 6.35 | < .001 |
| SAPS: Item total<sup>b</sup> | 12.51 (14.56) | - | - | - |
| Global summary<sup>b</sup> | 4.49 (4.15) | - | - | - |
| BNSS | 12.21 (12.09) | - | - | - |
| MADRS | 10.08 (9.11) | - | - | - |
| YMRS | 7.08 (6.75) | - | - | - |
| MCAS | 48.33 (4.23) | - | - | - |
| CPZ Equivalent (mg) | 443.55 (526.72) | - | - | - |
| Duration of Illness (Years) | 12.37 (9.32) | - | - | - |

Notes: ASI3, Anxiety Sensitivity Index – 3(49); BMI, body mass index; BNSS, Brief Negative Symptom Scale; CPZ, chlorpromazine; HC, healthy controls; IAS, Interoceptive Accuracy Scale(50); IATS, Interoceptive Attention Scale(51); IPASE, Inventory of Psychotic-like Anomalous Self-Experiences(52); MADRS, Montgomery-Asberg Depression Rating Scale; MCAS, Multnomah Community Ability Scale; NAART, North American Adult Reading Test; SAPS, Scale for the Assessment of Positive Symptoms; SZ, participants with schizophrenia spectrum disorder; TAS-20, Toronto Alexithymia Scale(53); YMRS, Young Mania Rating Scale.
<sup>a</sup>Parental education level: 1 = < 7 years; 2 = 7-9 years; 3 = 10-11 years; 4 = 12 years; 5 = 13-15 years; 6 = 16-17 years; 7 = 18+ years (graduate degree).
<sup>b</sup>Item total: sum of individual symptom ratings; Global summary: sum of global item ratings.

### Group Differences in Heartrate Changes

Full results of all multilevel models can be found in supplementary tables S1 – S6. For the model predicting heartrate changes using IAPS normative ratings (Model 1), here we focus on significant three-way interactions involving group only (see supplemental results for other significant effects). We found two significant three-way interaction effects: time × group × normative arousal, *p* = .019; and time^2^ × group × normative valence, *p* = .016. To understand the three-way interaction effects, we calculated estimates based on low (1 SD below the mean) and high (1 SD above the mean) IAPS normative arousal and valence ratings. We found that the initial IBI increase in HC was moderated by the normative arousal rating such that the higher the rating, the larger the change (Figure 2). In other words, HC exhibited a larger decrease in heartrate when viewing normatively high-arousal images relative to low-arousal ones. Similarly, the later IBI decrease in HC was moderated by the IAPS normative valence ratings such that the higher the rating, the faster the decrease (Figure 3). In other words, HC exhibited a faster return to baseline heartrate when viewing normatively pleasant images relative to unpleasant ones. But in SZ, both the time × normative arousal (*p* = .97) and the time^2^ × normative valence (*p* = .32) interactions were not significant. In other words, SZ exhibited no differences in heartrate changes across images of different normative arousal and valence ratings.

**Figure 2.**
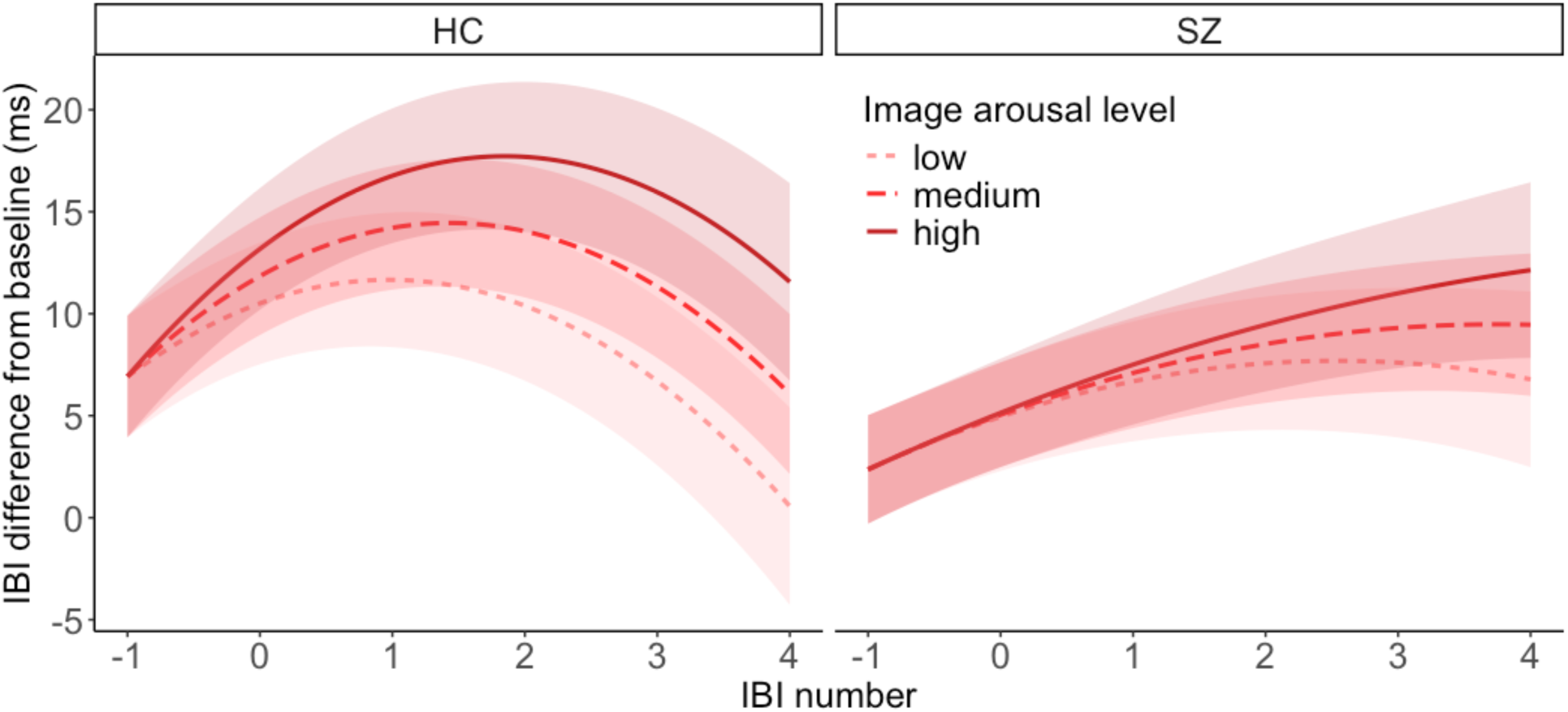
Time × group × normative arousal level interaction. Shading represents 95% confidence interval. IBI, inter-beat-intervals (the time difference between two consecutive R-peaks); HC, healthy controls; SZ, participants with schizophrenia spectrum disorder.

**Figure 3.**
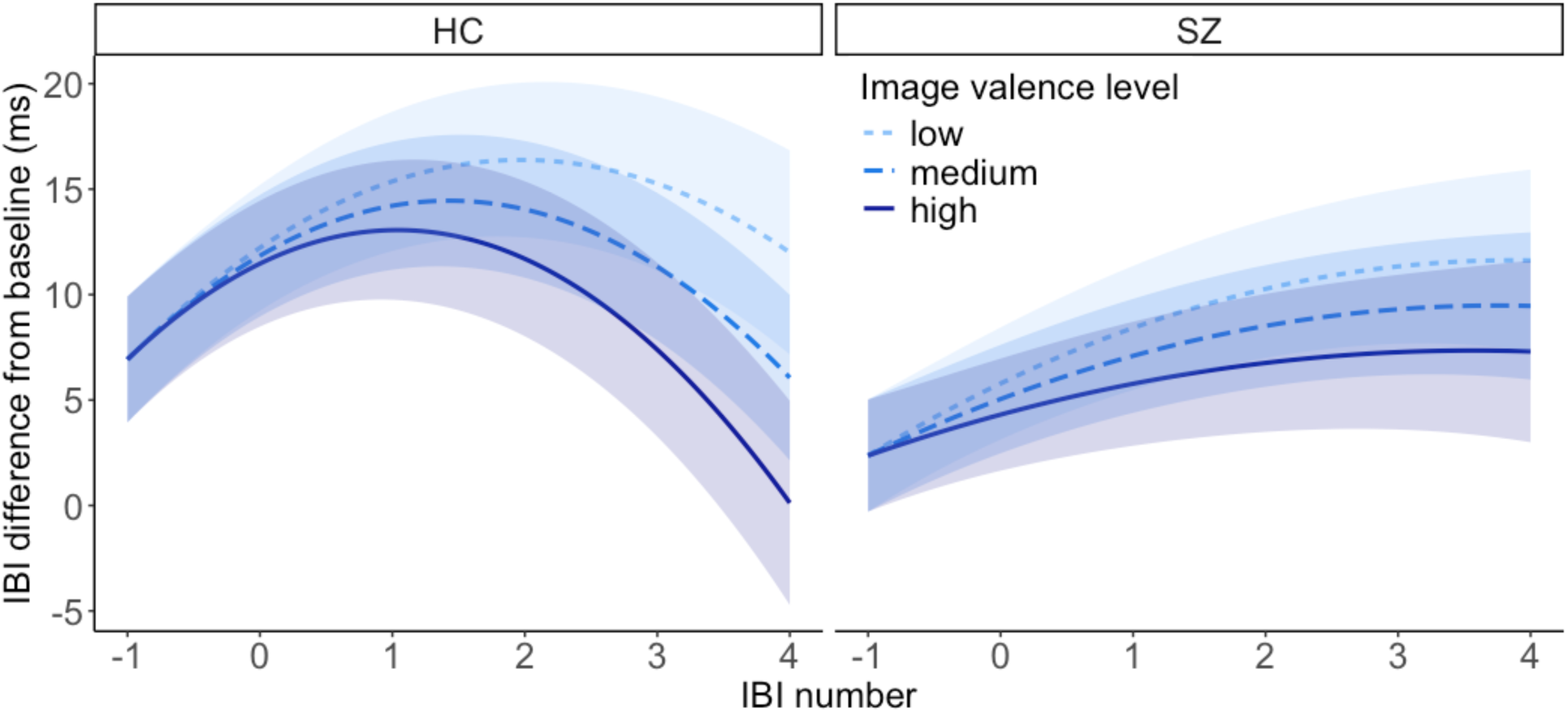
Time × group × normative valence level interaction. Shading represents 95% confidence interval. IBI, inter-beat-intervals (the time difference between two consecutive R-peaks); HC, healthy controls; SZ, participants with schizophrenia spectrum disorder.

For the model predicting heartrate changes using participants’ subjective ratings (Model 2), here we focus on interaction effects involving subjective ratings only, as the other effects are identical to Model 1. We found a significant three-way interaction effect: time × group × subjective arousal, *p* = .013. Follow-up analysis revealed that the time × subjective arousal interaction was significant in HC (*p* = .004), but not in SZ (*p* = .61). Specifically, we found that the higher the subjective arousal rating was, the larger the initial IBI increase in HC was (Figure 4). In other words, HC exhibited a larger decrease in heartrate when viewing subjectively high-arousal images relative to low-arousal ones. But SZ exhibited no differences in heartrate changes across images of different subjective arousal ratings.

**Figure 4.**
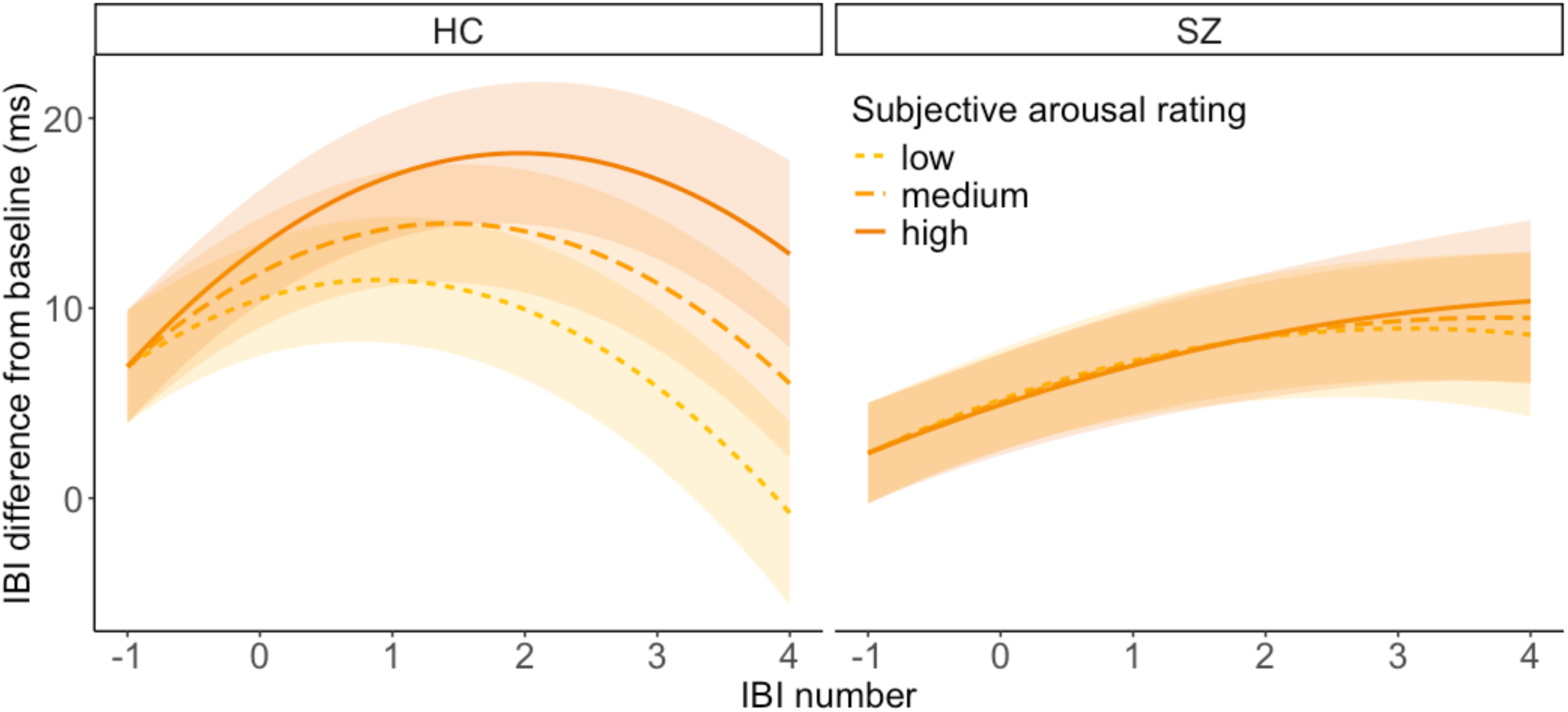
Time × group × subjective arousal rating interaction. Shading represents 95% confidence interval. IBI, inter-beat-intervals (the time difference between two consecutive R-peaks); HC, healthy controls; SZ, participants with schizophrenia spectrum disorder.

### Group Differences in HEP amplitude

We found a significant group × condition interaction effect in C-R, *F*(1,64) = 5.11, *p* = .027 (Figure 5). Follow-up analysis revealed that the effect of condition was significant in both groups, HC: *F*(1,30) = 46.2, *p* < .001, SZ: *F*(1,34) = 67, *p* < .001. But the effect of group was significant only in the LA condition, *F*(1,64) = 5.66, *p* = .02, and not in the HA condition, *F*(1,64) = 1.41, *p* = .24. More specifically, SZ showed a less negative peak voltage (i.e., a smaller peak because LA HEP was scored as the lowest negative peak; *M* = −0.48, *SD* = 0.49) than HC (*M* = −0.80, *SD* = 0.61; see Figure S3 for grand average waveforms and topographies). See supplemental results for main effects of condition.

**Figure 5.**
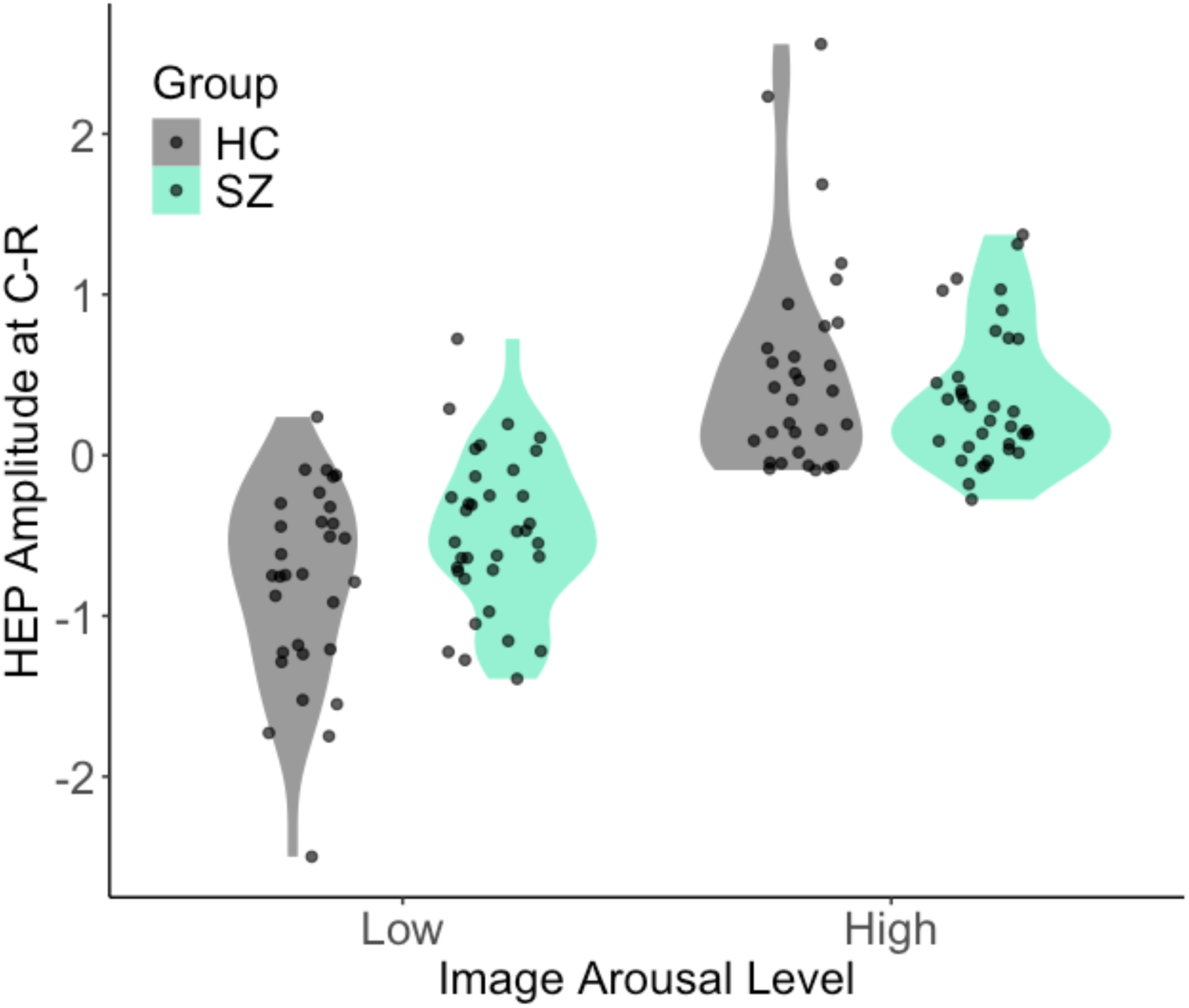
Group × condition interaction effect at C-R. HEP, heartbeat-evoked potential; C-R, mean of C2, C4, & C6 channels; HC, healthy controls; SZ, participants with schizophrenia spectrum disorder.

### Relationship between heartrate changes and HEP amplitude

For the model predicting heartrate changes using participants’ HEP amplitude at C-R, here we focus on interaction effects involving HEP amplitude only, as effects involving only time and group are similar to Model 1. We did not find any significant main or interaction effects involving HEP amplitude, suggesting that differences in HEP amplitude were not directly associated with changes in cardiac dynamics, and this did not differ between the two groups.

### Moderation Effects of Symptom Severity

For the model examining the effect of positive symptom severity on patterns of heartrate changes, we found two significant interaction effects: time × SAPS, *p* = .0098; and time^2^ × SAPS, *p* < .001. Follow-up analyses revealed that across all images, SZ with less severe positive symptoms exhibited both a larger initial increase in IBI, *b* = 4.10, *p* < .001, and a larger decrease in IBI after, *b* = −0.57, *p* < .001. While those with more severe positive symptoms exhibited a smaller initial increase in IBI, *b* = 1.99, *p* < .001, and a negligible decrease in IBI after, *b* = −0.07, *p* = .66. In other words, patients with less severe positive symptoms exhibited a pattern of IBI change that was more similar to that observed in HC. However, this effect was not moderated by their subjective ratings of either arousal or valence.

For the model examining the effect of negative symptom severity on patterns of heartrate changes, we found four significant interaction effects: time × BNSS × subjective arousal, *p* = .02; time^2^ × BNSS × subjective arousal, *p* = .029; time × BNSS × subjective valence, *p* = .003; and time^2^ × BNSS × subjective valence, *p* = .001. Here we only focus on the interaction with time × subjective arousal rating effect because that was the only significant interaction effect in Model 2. Follow-up analyses revealed that SZ with less severe negative symptoms exhibited a moderation effect of subjective arousal rating on IBI change that was more similar to that observed in HC (i.e., a larger initial decrease in heartrate when viewing subjectively high-arousal images relative to low-arousal ones). Exploratory analyses found similar effects when modeling the severity of avolition specifically (see supplemental results).

## Discussion

This study extends prior HEP research by testing arousal-related HEP modulation during emotional image viewing, a paradigm design that has not previously been applied to schizophrenia. Using concurrent self-report, ECG, and EEG measures, we examined peripheral and central markers of interoception and their associations with subjective emotional experience in people with schizophrenia spectrum disorders and healthy controls. We found that images with more intense emotional content led to larger momentary heartrate decelerations and higher subjective arousal ratings in healthy controls, but did not modulate heartrate changes in schizophrenia group and were less predictive of their subjective ratings. Similarly, subjective arousal ratings were predictive of cardiac dynamics in healthy controls, but not in schizophrenia participants. Moreover, we found smaller HEP amplitude in people with schizophrenia relative to healthy controls, which was unrelated to the group differences in cardiac dynamics. Lastly, the pattern of heartrate changes was moderated by both positive and negative symptoms in people with schizophrenia, such that those with less symptom burdens exhibited more normative cardiac dynamics. These results suggest a complex picture of both reduced cardiac reactivity to emotional stimuli and a disconnect between interoceptive afferent signals and cortical processing, leading to differentially reported subjective emotional experiences in people with schizophrenia.

Current findings add to the literature of emotional differences in SZ and suggest altered interoception as a potential underlying mechanism. We were able to replicate previous findings of different emotional arousal in SZ indexed by both subjective reports (10) and heartrate changes (13,36,37). Moreover, because we recorded subjective and physiological measures at the same time, we were able to directly assess the correspondence between the two within the same individual. Our findings revealed that the widely replicated finding of altered arousal report in SZ was not merely a response bias (i.e., a bias solely in the interpretation of bodily signals). Instead, there seems to be disruptions in both peripheral and central interoception processes. Overall, we found a restricted range in both heartrate changes and subjective ratings in SZ when viewing emotional images. While the image intensity modulated cardiac reactivity in HC, it had no effect on the heartrate changes in SZ, suggesting altered peripheral interoceptive signals. On the other hand, the cortical processing of heartbeat signals in SZ differed from HC, especially when viewing less intense images. Importantly, we found no relationship between heartrate changes and HEP amplitude in either group, suggesting that the changes in HEP amplitude were not merely a result of changes in cardiac dynamics (14), but do reflect genuine changes in central interoceptive processes. Therefore, the smaller HEP amplitude in SZ suggests altered interoception - a disconnect between central processing and afferent signals. This interpretation is further supported by the finding of a lack of correspondence between heartrate changes and subjective arousal ratings in SZ. In other words, the heartrate changes were not informative when SZ reported on their subjective emotional experience. Taken together, these findings suggest disruptions in both peripheral and central interoceptive processes in SZ that may have direct implications for emotional experience.

Interestingly, we found that both positive and negative symptoms modulated the pattern of heartrate changes in SZ, but in slightly different ways. Positive symptoms moderated cardiac dynamics, such that those with less severe symptoms exhibited more dynamic heartrate changes in general. However, there was no interaction effect between positive symptoms and subjective emotional ratings. It is possible that this reflects an overall change in cardiac dynamics from antipsychotic use. However, the impact of antipsychotic medication on baseline cardiac dynamics is typically small and non-significant (38), and we did not find a correlation between CPZ equivalent and baseline heartrate or HEP amplitude in the current sample either. On the other hand, we found that in SZ with less severe negative symptoms, subjective arousal rating modulated their heartrate changes in a more normative manner. This suggests that cardiac interoception may be more directly implicated in negative symptom manifestation. However, evidence has been mixed in clinical correlates of cardiac interoception alterations in SZ (15–17,39–43), so it is important to replicate current findings in future research. In sum, these findings suggest that disrupted interoception may be an underlying mechanism of schizophrenia symptoms (5), especially alterations in motivational and affective functioning.

The current study has a few limitations. First, because the majority of SZ were taking antipsychotic medications, the current findings could be confounded by potential medication effects. Though we did not find a correlation between CPZ equivalent and baseline heartrate or HEP amplitude, it is possible that antipsychotic (and other psychotropic) medications may have unintended effects on cardiac dynamics during states of high arousal only. Future research could benefit from studying medication-naïve patients or people at clinical high risk for psychosis. It may also be worthwhile to systematically examine the effects of antipsychotic medication during autonomic challenges. Relatedly, the two groups in the current study were not matched on baseline heartrate, which may limit the generalizability of the findings. It is not uncommon to find an increased baseline heartrate in people with schizophrenia (39,44,45), but it is unclear whether this is a result of medication and other secondary factors, or part of the illness phenotype. Moreover, the current finding of restricted cardiac reactivity to emotional images is perhaps in line with the larger literature of reduced baseline heartrate variability in people with schizophrenia (46), but it is unclear whether these come from disturbances in a shared mechanism or separate ones. Future research should examine cardiac dynamics and heart-brain interactions under a wider range of conditions. Next, there is considerable heterogeneity in the methods to compute HEP in the current literature (14,47,48). The lack of consensus in a standard procedure makes it difficult to directly compare current findings with existing literature. Future research on standardizing the methodology is crucial to move the field forward. Relatedly, we did not correct for multiple testing when examining group differences in HEP amplitude. Because our effect of interest was an interaction effect, which tend to have smaller effect sizes, we decided it was more important to preserve statistical power and protect against potential Type II errors. Nevertheless, it is important to replicate current findings with larger samples in the future. Lastly, the current group of participants were only experiencing minimal psychotic symptoms in the somatic and passivity domains – areas that theoretically have the closest connection with interoception; a more enriched sample may allow us to identify more specific relationships between interoception alterations and clinical symptoms.

In conclusion, we found evidence of alterations in both peripheral and central interoception in people with schizophrenia, especially in those with more severe symptoms. Current study demonstrates the utility of multimodal data in bridging biological and subjective levels of understanding. These findings highlight the importance of interoception when studying subjective emotional experience and illness mechanisms of schizophrenia.

## Data Availability

All data produced in the present study are available upon reasonable request to the authors.

## Acknowledgments

This work was supported in part by the Harvard Medical School Department of Psychiatry (Livingston Award to BY), the McLean Hospital (Presidential Award to BY), and philanthropic gift and divisional funds in the Psychotic Disorders Division at McLean Hospital. The funding sources had no involvement in the study design, data collection, analysis, and interpretation, and the writing of and decision to submit the article for publication. The authors would also like to thank all participants for their participation in this study.

