## Supplement for "Altered interoception in schizophrenia and its role in subjective emotional experience"

### Supplemental Methods

#### Participants

Exclusion criteria for all participants included a history of significant medical or neurological illness that actively interferes with brain or cognitive functioning (e.g., epilepsy) or affects peripheral nervous system and/or bodily sensations (e.g., diabetes mellitus, heart arrhythmia with frequent symptoms), head injury with loss of consciousness > 1 hour, electroconvulsive therapy or transcranial magnetic stimulation within 6 months prior to study entry, DSM-5 severe substance use disorder within 90 days prior to study entry, and vision that was not (corrected to) normal. Healthy controls (HC) were additionally excluded for personal or first-degree familial history of bipolar or schizophrenia spectrum disorders, personal history of other major psychiatric diagnoses (any history of anxiety or PTSD must be < 1 year duration, remitted for > 1 year, and did not require medication therapy), psychiatric hospitalization, or psychotropic use within 3 months prior to study entry.

#### Assessments

All participants completed the following self-report scales measuring various constructs related to interoception: the Interoceptive Accuracy Scale (IAS)(1) measures one's belief of ability to accurately perceive the state of one's body; the Interoceptive Attention Scale (IATS)(2) measures the propensity of focusing one's attention on internal signals; the Anxiety Sensitivity Index – 3 (ASI3)(3) measures fear of arousal-related sensations; the Twenty-item Toronto Alexithymia Scale (TAS-20)(4) measures alexithymia (difficulty understanding one's own emotions); the Inventory of Psychotic-like Anomalous Self-Experiences (IPASE)(5) measures a wide range of disturbances in the subjective experience of self.

#### Task and Procedure

All photo stimuli were converted to 8-bit grayscale(6) and modified using GIMP (version 2.10.32) so that their brightness and contrast were identical. All photos are landscape in orientation (1024 × 768) and contain human. Images were grouped into four runs of 24, with 8 photos of each valence (pleasant, neutral, and unpleasant) in each run. The order of images was pseudo-randomized so that no more than 2 photos of the same valence were presented consecutively. Before beginning the task, participants completed 3 practice trials to get familiarized with the task instructions and the self-assessment manikin (SAM) rating procedure. There was no time limit on response, but participants were encouraged to rate according to their initial reaction.

#### Apparatus and Setup

Participants sat in a dimly lit room, at a distance of approximately 62 cm in front of a computer screen (screen size: 52.6 × 29.6 cm; resolution: 1920 × 1080). Participants used a computer mouse to respond. We used Presentation (Neurobehavioral Systems, San Francisco, CA) to present the stimuli and record responses. Electroencephalogram (EEG) signals were recorded continuously online using the BioSemi Active Two system (BioSemi Inc, Amsterdam, The Netherland) from 64 Ag-AgCl electrodes fitted in a stretch Lycra cap, at a sampling rate of 1024 Hz, with a bandpass of DC-104 Hz. The Common Mode Sense (CMS) active electrode (PO1 site) and Driven Right Leg (DRL) passive electrode (PO2 site) served as the ground for online reference for all other electrodes. Two electrodes were placed on the left outer canthi (HEOG) and below the left eye (VEOG) to monitor blinks and eye movements. Two electrodes were placed on the left and right mastoids so that the EEG data could be re-referenced offline to the averaged mastoid. Electrocardiogram (ECG) signals were recorded continuously using the same BioSemi Active Two system from two electrodes placed over the right clavicle and left iliac crest(7).

**ECG & EEG Preprocessing**

ECG and EEG data were down sampled to 512 Hz and processed offline using BrainVision Analyzer 2.2 (Brain Products, Germany). Channels with overall poor signal quality (< 50% usable data) were excluded from further processing. We applied a bandpass filter of 0.5 – 40 Hz(8) and segmented the data using a window of -3.5 to 6 s (i.e., from 3.5 s before stimulus onset to stimulus offset), to ensure at least two R-peaks before stimulus onset were retained for baseline heartrate calculation. The epochs were baseline corrected using a window from 200 ms before stimulus onset to stimulus onset. R-peaks in the ECG trace were automatically identified using the Analyzer built-in function. We then visually inspected all identified R-peaks and made corrections when necessary. We applied standard Independent Component Analysis (ICA) and inverse ICA to EEG data for artifact correction. All ICA components were visually inspected to identify and remove artifacts from ocular, muscle, cardiac, and single-channel noises. We then applied the Analyzer built-in function to correct pulse-related movement artifacts in all channels.

For heartbeat-evoked potential (HEP) analysis, we segmented the epochs to 150 – 6000 ms after stimulus onset to avoid including activities related to initial visual processing of the images. Within these windows, we then segmented the data to -125 - 600 ms around each R-peak, and baseline corrected using a window of 125 - 25 ms before R-peak to avoid including the start of the R-wave(8). Epochs with remaining artifacts were identified and rejected automatically using the following criteria: exceeding a voltage step of 50 μV/ms, exceeding the max/min difference of 200 μV/ms, amplitude < -100 μV or > 100 μV, and activity within the epoch < 0.5 μV. Remaining data were then smoothed by using a moving average window of 7 data points, then averaged within each channel for peak identification. On average, HC participants had 181 Low Arousal (LA) epochs and 361 High Arousal (HA) epochs, while participants with schizophrenia-spectrum disorders (SZ) had 218 LA epochs and 440 HA epochs. There are no group or condition differences in minimal percentage of usable epochs out of all channels (HC: 96.7% LA epochs, 97.2 % HA epochs; SZ: 96.2% LA epochs, 96.6% HA epochs).

**Statistical Analysis**

For the multilevel linear regression models, group was coded as 0 = HC and 1 = SZ. All continuous predictors were grand mean centered. For significant interaction effects, we followed up with simple slope analysis(9) and/or by rerunning the model with the opposite coding of the group variable (i.e., 0 = SZ and 1 = HC) to obtain parameter estimates for SZ.

**Subjective ratings.** To assess the correspondence between IAPS normative arousal ratings and participants' subjective arousal ratings, we constructed a multilevel model predicting subjective rating on each trial using IAPS normative rating, group, and normative rating × group interaction term. To account for individual differences in subjective ratings generally and in the effect of normative ratings, we included a random intercept for subject and a random slope for normative rating. To account for potential rating differences due to specific image content, we also included a random intercept for trial.

**Heartrate changes.** The final model predicting the effect of IAPS images on heartrate changes (Model 1) included the following predictors: time, $time^2$, group, time × group, $time^2$ × group, time × normative arousal, $time^2$ × normative arousal, time × normative valence, $time^2$ × normative valence, time × group × normative arousal, $time^2$ × group × normative arousal, time × group × normative valence, and $time^2$ × group × normative valence. Time was centered on IBI – 1.

**Relationship between heartrate changes and HEP amplitude.** The model predicting heartrate changes using HEP amplitude included the following predictors: time, $time^2$, group, HEP, time × group, $time^2$ × group, time × HEP, $time^2$ × HEP, time × group × HEP, and $time^2$ × group × HEP. To account for general inter-individual differences in heartrate changes, we included a random intercept for subject and a random slope of time for condition within subject.

**Symptom associations.** The model exploring the modulation effect of positive symptom severity on heartrate changes included the following predictors: time, $time^2$, time × SAPS, $time^2$ × SAPS, time × subjective arousal, $time^2$ × subjective arousal, time × subjective valence, $time^2$ × subjective valence, time × SAPS × subjective arousal, $time^2$ × SAPS × subjective arousal, time × SAPS × subjective valence, and $time^2$ × SAPS × subjective valence. In the negative symptom model, SAPS scores were substituted with BNSS scores. The random effects remained identical to Model 1.

We also explored associations between HEP amplitude and clinical symptom severity in the SZ group. For channels with a significant group × condition effect, we conducted Spearman's rank correlations between HEP amplitude and the total scores of SAPS, BNSS, ASI3, TAS-20, and IPASE, for LA and HA conditions separately.

### Supplemental Results

#### Effects of Age, Sex, BMI, and CPZ equivalent

For baseline heartrate, we found no correlations with age ($r$ = -.07, $p$ = .54) and BMI ($r$ = .21, $p$ = .09). There were no differences between males and females, $t$(68) = -0.89, $p$ = .38. In SZ, we found no correlation with CPZ equivalent, $r$ = .08, $p$ = .65.

We checked potential effects of age, sex, BMI, and CPZ equivalent on HEP amplitude at Fz, FCz, and Cz channels. For Fz, we found no correlation with age in LA ($r$ = .03, $p$ = .78) or HA ($r$ = .10, $p$ = .43) condition. We found no correlation with BMI in LA ($r$ = -.05, $p$ = .69) or HA ($r$ = -.10, $p$ = .42) condition. There were no differences between males and females in LA ($t$(48) = -0.76, $p$ = .45) or HA ($t$(28) = -1.10, $p$ = .28) condition. We found no correlation with CPZ equivalent in LA ($r$ = .19, $p$ = .28) or HA ($r$ = -.08, $p$ = .63) condition in SZ. For FCz, we found no correlation with age in LA ($r$ = -.04, $p$ = .72) or HA ($r$ = .24, $p$ = .05) condition. We found no correlation with BMI in LA ($r$ = -.05, $p$ = .69) or HA ($r$ = -.07, $p$ = .56) condition. There were no differences between males and females in LA ($t$(43) = -0.53, $p$ = .60) or HA ($t$(30) = -0.91, $p$ = .37) condition. We found no correlation with CPZ equivalent in LA ($r$ = .15, $p$ = .40) or HA ($r$ = -.10, $p$ = .56) condition. For Cz, we found no correlation with age in LA ($r$ = -.07, $p$ = .60) or HA ($r$ = .22, $p$ = .08) condition. We found no correlation with BMI in LA ($r$ = .04, $p$ = .76) or HA ($r$ = -.15, $p$ = .25) condition. There were no differences between males and females in LA ($t$(50) = -0.54, $p$ = .59) or HA ($t$(32) = -0.76, $p$ = .45) condition. We found no correlation with CPZ equivalent in LA ($r$ = .17, $p$ = .33) or HA ($r$ = -.16, $p$ = .37) condition.

#### Correspondence between Normative and Subjective ratings

We found a significant main effect of normative rating, $p < .001$ on subjective rating. As normative arousal rating increased, HC's subjective arousal rating also increased, $b = 0.83$, $p < .001$. We also found a significant normative rating × group interaction effect, $p = .026$. Follow-up analysis revealed that the effect of normative arousal rating was also statistically significant in SZ but smaller, $b = 0.59$, $p < .001$. In other words, the subjective rating was more closely aligned with the normative rating in HC than in SZ (Figure S2).

**Group Differences in Heartrate Changes**

For the model predicting heartrate changes using IAPS normative ratings (Model 1), we found significant main effects of time, $p < .001$; time$^2$, $p < .001$; and group, $p = .028$. We also found three significant two-way interaction effects: time × group, $p < .001$; time$^2$ × group, $p < .001$; and time × normative arousal, $p = .022$. Follow-up analyses revealed that across all images, the IBI in HC increases initially after stimulus onset, $b = 6.19$, $p < .001$, and this increase slows down over time, $b = -1.27$, $p < .001$. In other words, there was an overall decrease in heartrate after stimulus onset, followed by a return to baseline heartrate. The IBI change in SZ follows a similar pattern, but the initial increase is smaller than in HC, $b = 2.99$, $p < .001$, and the slowdown in increase is likewise smaller, $b = -0.32$, $p < .001$. Lastly, there was also a significant group difference in the relative change in IBI at time 0 (IBI – 1), $b = -4.56$, $p = .028$. Follow-up analysis revealed that there was a small increase in IBI at time 0 in HC (6.93, $p < .001$), but not in SZ ($p = .\ 085$).

**Condition differences in HEP amplitude**

We found a significant main effect of condition in all channels, Fz: $F(1,64) = 107.59$, $p < .001$, FCz: $F(1,64) = 108.55$, $p < .001$, Cz: $F(1,63) = 79.15$, $p < .001$, F-L: $F(1,64) = 96.42$, $p < .001$, F-R: $F(1,64) = 129.16$, $p < .001$, FC-L: $F(1,64) = 90.46$, $p < .001$, FC-R: $F(1,64) = 129.85$, $p < .001$, C-L: $F(1,64) = 88.38$, $p < .001$, C-R: $F(1,64) = 103.19$, $p < .001$. This effect was expected as HEP peak amplitude was mostly positive in HA and mostly negative in LA condition.

**Moderation Effects of Avolition**

For the model examining the effect of avolition severity (measured by BNSS avolition subscale) on patterns of heartrate changes, we found four significant interaction effects: time × avolition × subjective arousal, $p = .02$; time$^2$ × avolition × subjective arousal, $p = .01$; time × avolition × subjective valence, $p = .02$; and time$^2$ × avolition × subjective valence, $p = .002$. See table S7 for full model results. Here we only focus on the interaction with time × subjective arousal rating effect because that was the only significant interaction effect in Model 2. To better understand these interaction effects, we calculated estimates based on low (1 SD below the mean) and high (1 SD above the mean) avolition scores. Results revealed that participants with less severe avolition exhibited a moderation effect of subjective arousal rating on IBI change that was more similar to that observed in HC (i.e., a larger decrease in heartrate when viewing subjectively high-arousal images relative to low-arousal ones).

**Associations between HEP and clinical symptom severity**

We found a significant correlation between HEP amplitude at C-R and anxiety sensitivity (ASI3), $\rho = -0.35$, $p = .04$, and alexithymia (TAS-20), $\rho = -0.39$, $p = .02$, in the HA condition only. In other words, SZ with a smaller HEP amplitude at C-R on HA trials reported more fear of arousal-related sensations (Figure S4) and more difficulty understanding their own emotions (Figure S5).

**Table S1.** Multilevel regression coefficients estimating effects of group and normative arousal rating on subjective arousal rating.

| Variable | *b* | Standard Error | *F(df)* |
|---|---|---|---|
| Intercept | 5.2 | 0.21 | |
| SZ | 0.06 | 0.27 | 0.05 (68) |
| Normative arousal | 0.83 | 0.08 | 143.83*** (103) |
| SZ × normative arousal | -0.24 | 0.1 | 5.17* (68) |

Notes: SZ, participants with schizophrenia spectrum disorder.
**p* < .05, ***p* < .01, *** *p* < .001.

**Table S2.** Multilevel regression coefficients estimating effects of group, normative ratings, and time on heartrate changes.

| Variable | *b* | Standard Error | *F(df)* |
|---|---|---|---|
| Intercept | 6.93 | 1.52 | |
| SZ | -4.56 | 2.04 | 5.00* (79) |
| Time | 6.19 | 0.61 | 125.53*** (39665) |
| Time$^2$ | -1.27 | 0.11 | 124.08*** (33245) |
| SZ × Time | -3.19 | 0.82 | 15.18*** (39665) |
| SZ × Time$^2$ | 0.96 | 0.14 | 45.11*** (33245) |
| Normative arousal × Time | 1.05 | 0.34 | 5.22* (29801) |
| Normative arousal × Time$^2$ | -0.04 | 0.07 | 0.20 (33245) |
| Normative valence × Time | -0.09 | 0.23 | 2.68 (29801) |
| Normative valence × Time$^2$ | -0.1 | 0.04 | 1.20 (33245) |
| SZ × Normative arousal × Time | -1.06 | 0.45 | 5.46* (29801) |
| SZ × Normative arousal × Time$^2$ | 0.13 | 0.09 | 2.12 (33245) |
| SZ × Normative valence × Time | -0.33 | 0.31 | 1.14 (29801) |
| SZ × Normative valence × Time$^2$ | 0.14 | 0.06 | 5.84* (33245) |

**p* < .05, ***p* < .01, *** *p* < .001.

**Table S3.** Multilevel regression coefficients estimating effects of group, subjective ratings, and time on heartrate changes.

| Variable | *b* | Standard Error | *F(df)* |
|---|---|---|---|
| Intercept | 6.93 | 1.52 | |
| SZ | -4.56 | 2.03 | 5.03* (79) |
| Time | 6.2 | 0.61 | 125.84*** (39665) |
| Time$^2$ | -1.28 | 0.11 | 124.50*** (33246) |
| SZ × Time | -3.2 | 0.82 | 15.26*** (39665) |
| SZ × Time$^2$ | 0.96 | 0.14 | 45.41*** (33246) |
| Subjective arousal × Time | 0.65 | 0.23 | 3.26 (30022) |
| Subjective arousal × Time$^2$ | -0.002 | 0.04 | 0.38 (33346) |
| Subjective valence × Time | -0.22 | 0.22 | 0.59 (30114) |

| | | | |
|---|---|---|---|
| Subjective valence × Time² | -0.07 | 0.04 | 2.58 (33413) |
| SZ × Subjective arousal × Time | -0.75 | 0.3 | 6.15* (30022) |
| SZ × Subjective arousal × Time² | 0.04 | 0.06 | 0.47 (33346) |
| SZ × Subjective valence × Time | 0.21 | 0.3 | 0.50 (30114) |
| SZ × Subjective valence × Time² | 0.04 | 0.06 | 0.48 (33413) |

$^{*}p < .05$, $^{**}p < .01$, $^{***}\ p < .001$.

**Table S4.** Multilevel regression coefficients estimating effects of group, heartbeat-evoked potential (HEP) amplitude at C-R (the mean of C2, C4, and C6 sites), and time on heartrate changes.

| Variable | *b* | Standard Error | *F(df)* |
|---|---|---|---|
| Intercept | 7.13 | 1.56 | |
| SZ | -5.03 | 2.15 | 5.47* (77) |
| HEP | -0.93 | 0.68 | 1.86 (609) |
| Time | 5.6 | 0.85 | 56.25*** (560) |
| Time² | -1.21 | 0.13 | 75.93*** (577) |
| SZ × Time | -2.48 | 1.16 | 4.52* (561) |
| SZ × Time² | 0.85 | 0.18 | 22.26*** (577) |
| HEP × Time | 0.33 | 0.88 | 0.14 (501) |
| HEP × Time² | 0.13 | 0.14 | 1.20 (578) |
| SZ × HEP × Time | -0.28 | 1.3 | 0.05 (346) |
| SZ × HEP × Time² | 0.01 | 0.2 | 0.004 (598) |

$^{*}p < .05$, $^{**}p < .01$, $^{***}\ p < .001$.

**Table S5.** Multilevel regression coefficients estimating effects of positive symptom severity, subjective ratings, and time on heartrate changes.

| Variable | *b* | Standard Error | *F(df)* |
|---|---|---|---|
| Intercept | 2.37 | 1 | |
| Time | 3.05 | 0.43 | 51.20*** (22313) |
| Time² | -0.33 | 0.08 | 19.40*** (18570) |
| SAPS × Time | -0.26 | 0.1 | 6.67** (4659) |
| SAPS × Time² | 0.07 | 0.02 | 15.32*** (7171) |
| Subjective arousal × Time | -0.15 | 0.16 | 0.88 (16701) |
| Subjective arousal × Time² | 0.05 | 0.03 | 3.09 (18260) |
| Subjective valence × Time | -0.01 | 0.15 | 0.009 (17967) |
| Subjective valence × Time² | -0.02 | 0.03 | 0.59 (18665) |
| SAPS × Subjective arousal × Time | 0.05 | 0.04 | 1.22 (16531) |
| SAPS × Subjective arousal × Time² | -0.01 | 0.008 | 2.90 (18178) |
| SAPS × Subjective valence × Time | 0.05 | 0.04 | 2.07 (17937) |
| SAPS × Subjective valence × Time² | -0.01 | 0.007 | 2.06 (18645) |

Notes: SAPS, Scale for the Assessment of Positive Symptoms.
$^{*}p < .05$, $^{**}p < .01$, $^{***}\ p < .001$.

**Table S6.** Multilevel regression coefficients estimating effects of negative symptom severity, subjective ratings, and time on heartrate changes.

| Variable | *b* | Standard Error | *F(df)* |
|---|---|---|---|
| Intercept | 2.37 | 1.02 | |
| Time | 2.99 | 0.42 | 49.67*** (22312) |
| Time$^2$ | -0.31 | 0.07 | 17.66*** (18555) |
| BNSS × Time | 0.02 | 0.03 | 0.34 (4815) |
| BNSS × Time$^2$ | -0.006 | 0.006 | 1.15 (7333) |
| Subjective arousal × Time | -0.13 | 0.16 | 0.69 (17249) |
| Subjective arousal × Time$^2$ | 0.04 | 0.03 | 1.90 (18529) |
| Subjective valence × Time | 0.08 | 0.16 | 0.24 (17950) |
| Subjective valence × Time$^2$ | -0.05 | 0.03 | 2.19 (18651) |
| BNSS × Subjective arousal × Time | -0.03 | 0.01 | 5.28* (17787) |
| BNSS × Subjective arousal × Time$^2$ | 0.006 | 0.003 | 4.77* (18741) |
| BNSS × Subjective valence × Time | 0.04 | 0.01 | 8.63** (17940) |
| BNSS × Subjective valence × Time$^2$ | -0.009 | 0.003 | 10.79** (18645) |

Notes: BNSS, Brief Negative Symptom Scale.
$^*p < .05$, $^{**}p < .01$, $^{***}p < .001$.

**Table S7.** Multilevel regression coefficients estimating effects of avolition severity (indexed by BNSS subscale), subjective ratings, and time on heartrate changes.

| Variable | *b* | Standard Error | *F(df)* |
|---|---|---|---|
| Intercept | 2.37 | 1.02 | |
| Time | 3.01 | 0.42 | 50.50*** (22313) |
| Time$^2$ | -0.32 | 0.07 | 18.28*** (18558) |
| Avolition × Time | 0.29 | 0.19 | 2.32 (4803) |
| Avolition × Time$^2$ | -0.001 | 0.03 | 0.001 (7326) |
| Subjective arousal × Time | -0.19 | 0.16 | 1.45 (17446) |
| Subjective arousal × Time$^2$ | 0.06 | 0.03 | 3.25 (18606) |
| Subjective valence × Time | 0.07 | 0.16 | 0.19 (17975) |
| Subjective valence × Time$^2$ | -0.05 | 0.03 | 2.18 (18659) |
| Avolition × Subjective arousal × Time | -0.2 | 0.08 | 5.74* (17931) |
| Avolition × Subjective arousal × Time$^2$ | 0.04 | 0.02 | 6.41* (18777) |
| Avolition × Subjective valence × Time | 0.2 | 0.08 | 5.56* (17966) |
| Avolition × Subjective valence × Time$^2$ | -0.05 | 0.02 | 9.62** (18653) |

$^*p < .05$, $^{**}p < .01$, $^{***}p < .001$.

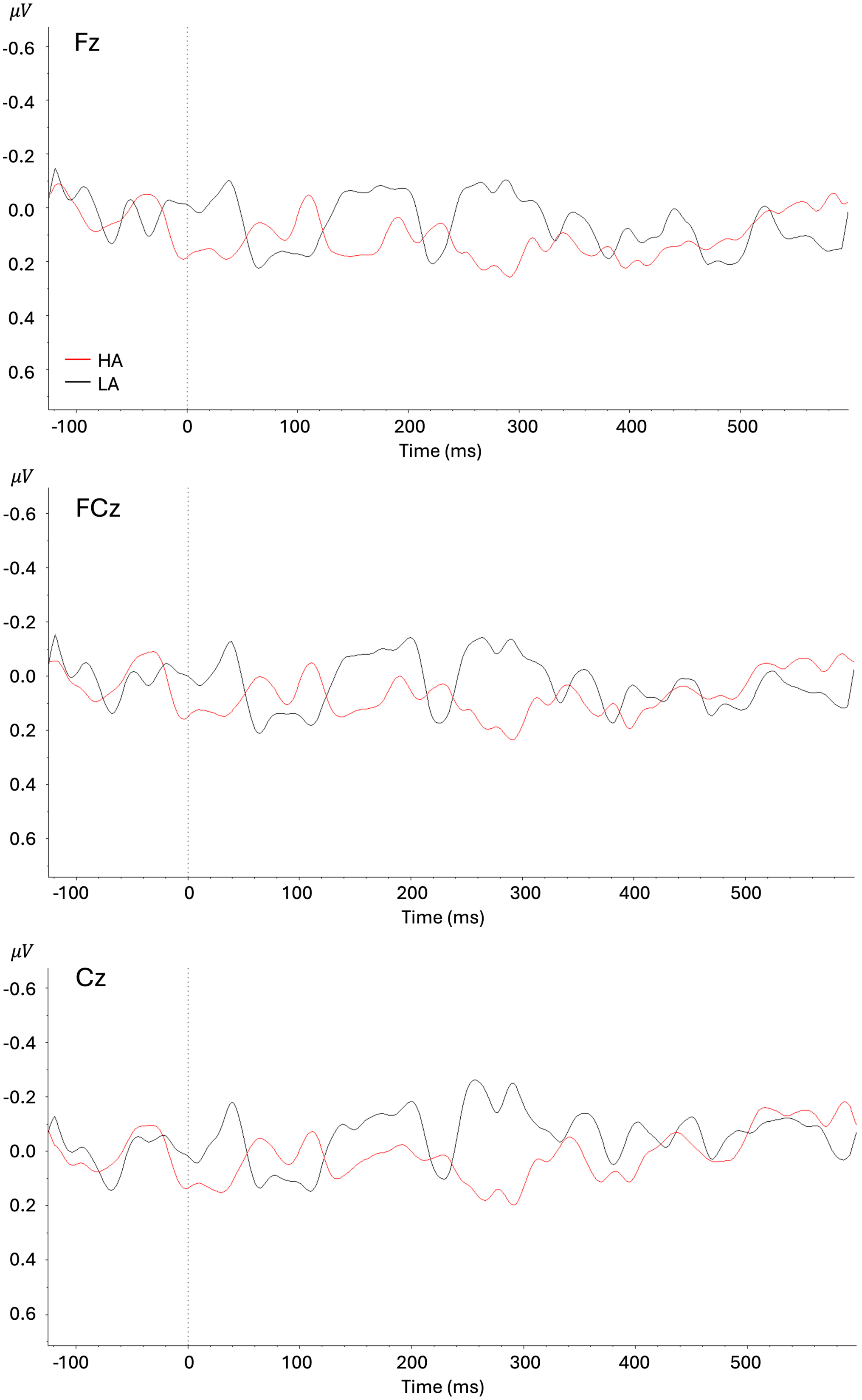


**Figure S1.** Grand average waveforms from healthy controls at Fz, FCz, and Cz channels. HA, high arousal; LA, low arousal.

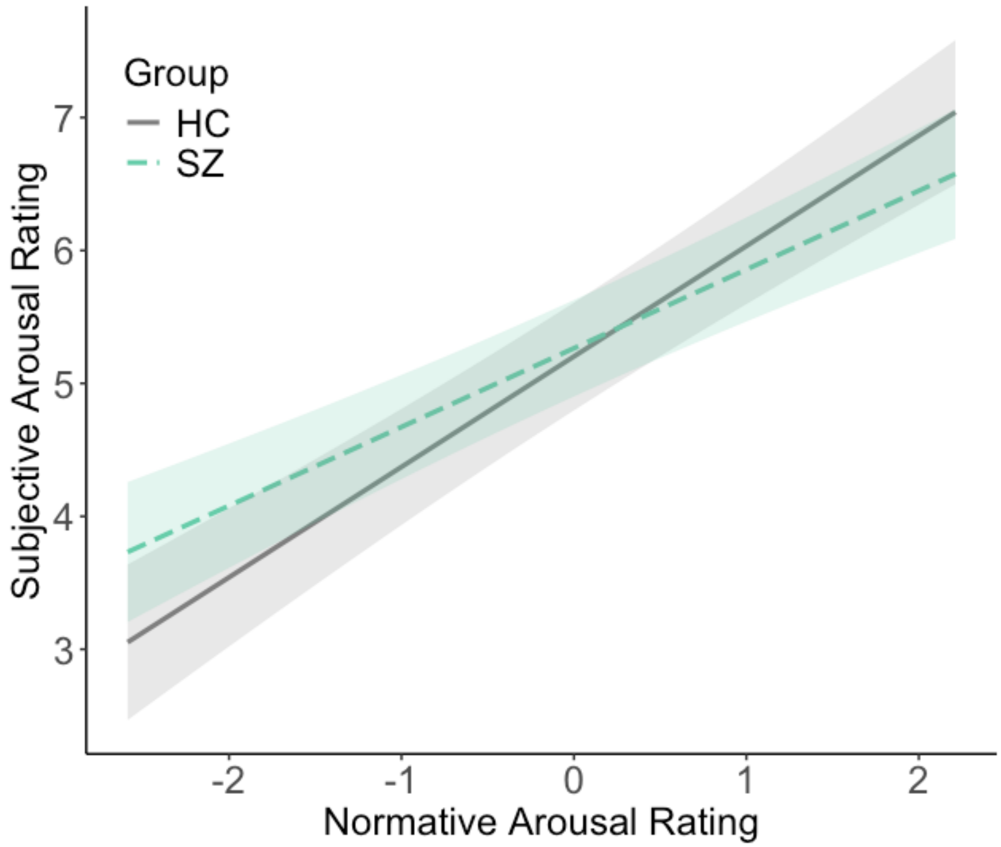


**Figure S2.** Normative rating × group interaction. Shading represents 95% confidence interval. HC, healthy controls; SZ, participants with schizophrenia spectrum disorder.

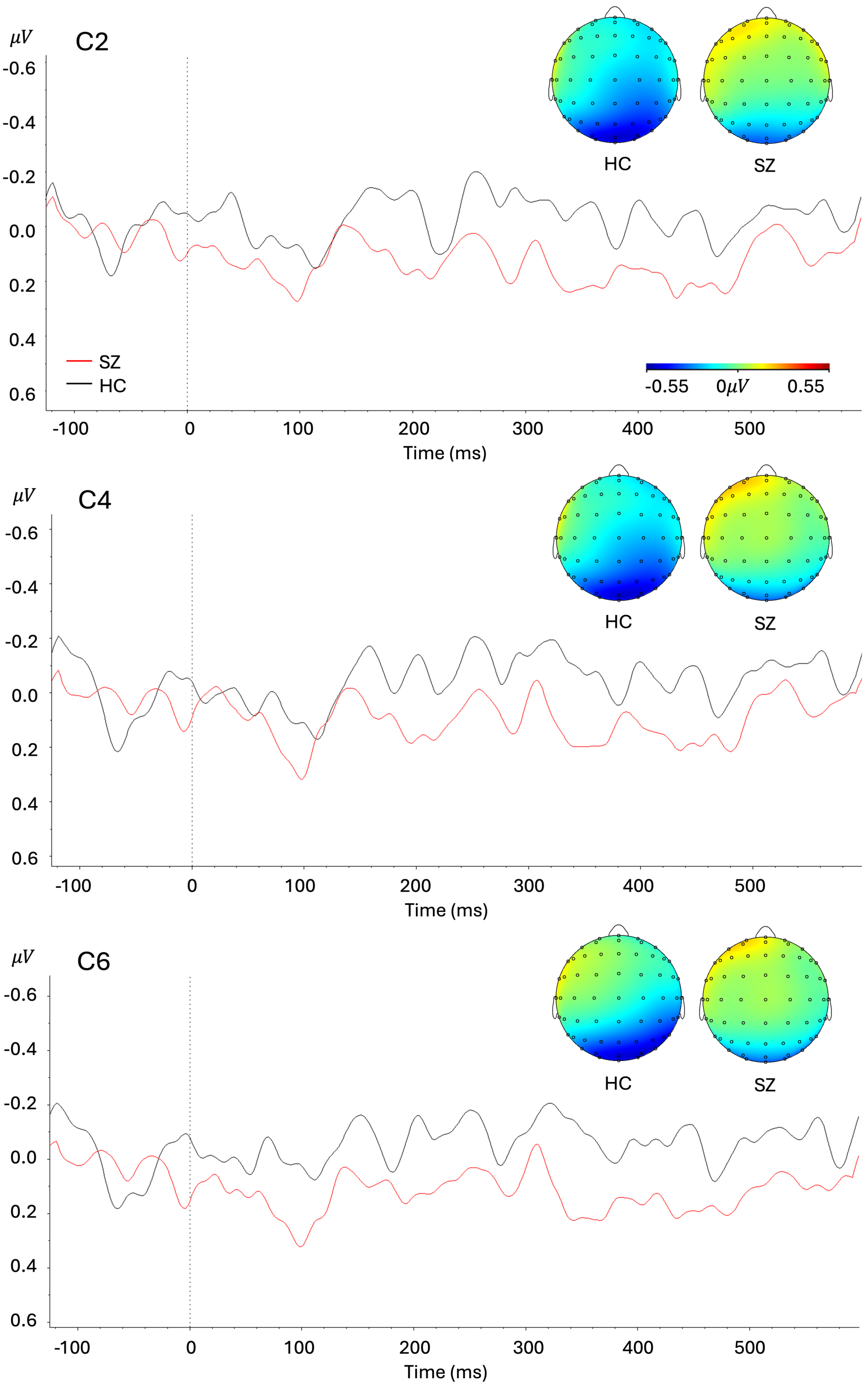


**Figure S3.** Grand average waveforms from the low arousal condition at C2, C4, and C6 channels, with topographies at the peak amplitude of heartbeat-evoked potential (HEP). HC, healthy controls; SZ, participants with schizophrenia spectrum disorder.

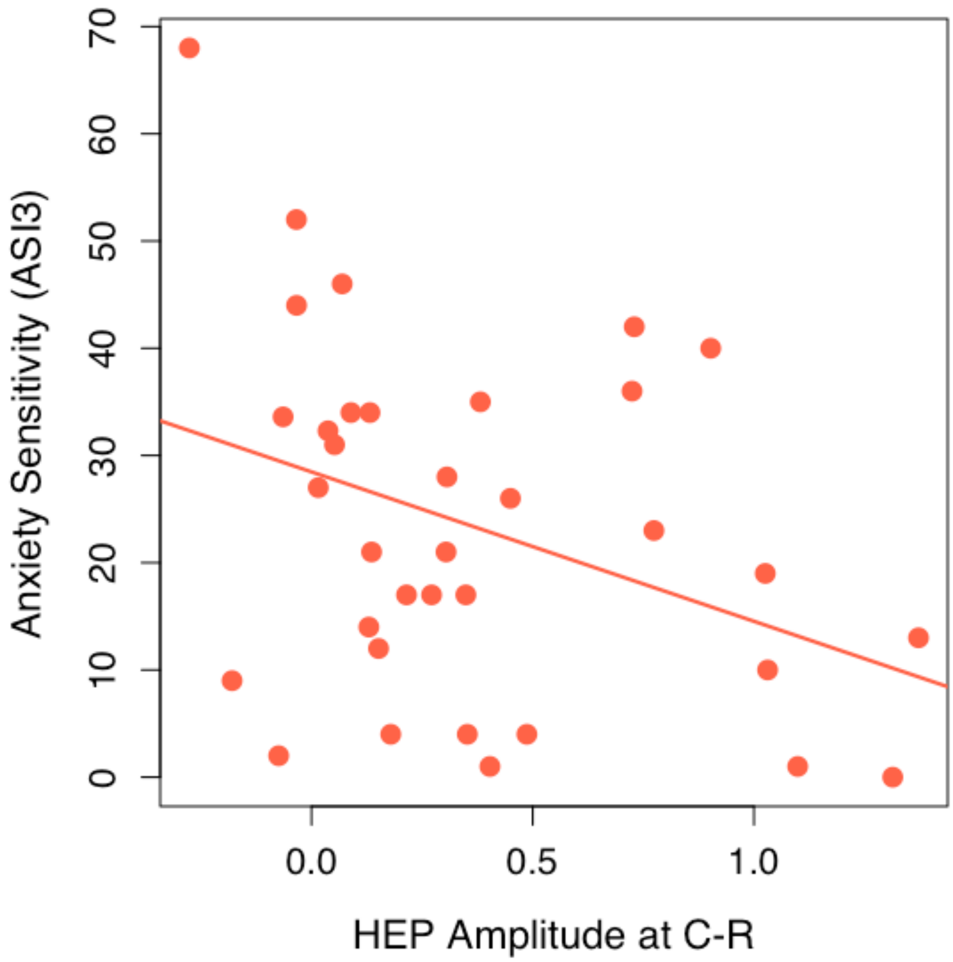


**Figure S4.** Correlation between anxiety sensitivity and HEP amplitude at C-R in high arousal condition in SZ. ASI3, Anxiety Sensitivity Index – 3; C-R, mean of C2, C4, & C6 channels; HEP, heartbeat-evoked potential; SZ, participants with schizophrenia spectrum disorder.

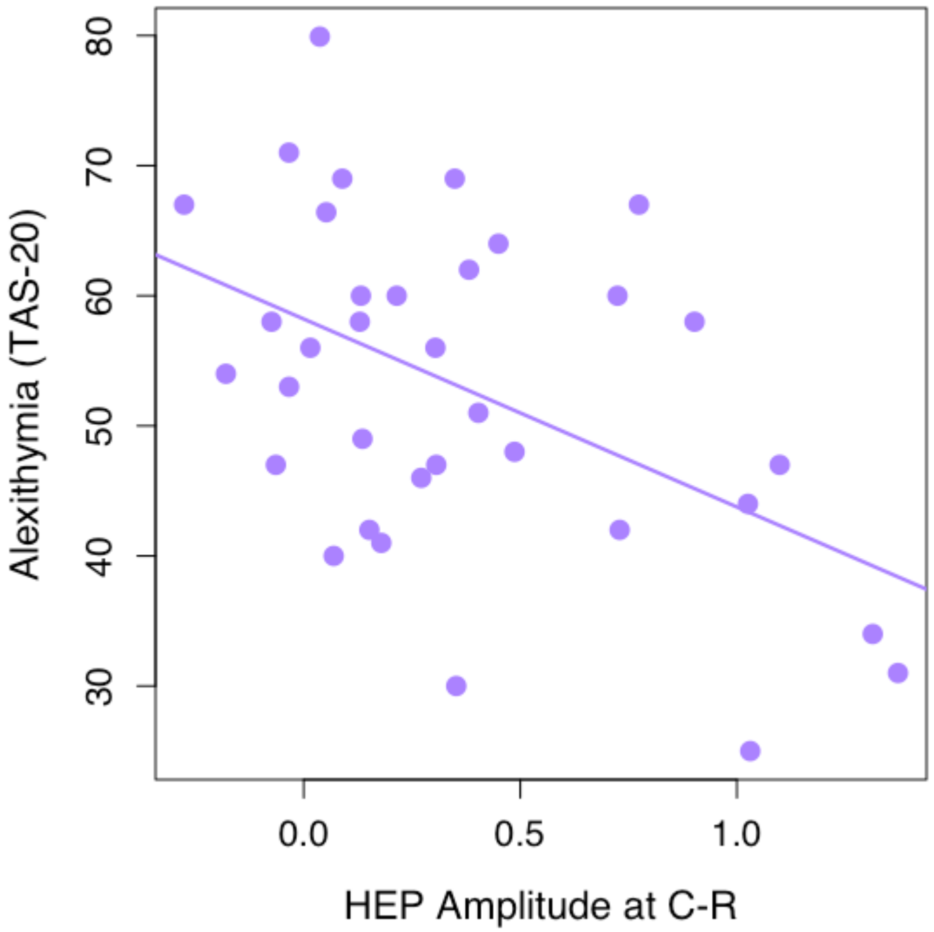


**Figure S5.** Correlation between alexithymia and HEP amplitude at C-R in high arousal condition in SZ. C-R, mean of C2, C4, & C6 channels; HEP, heartbeat-evoked potential; SZ, participants with schizophrenia spectrum disorder; TAS-20, Toronto Alexithymia Scale.
